# The Relationship Between Work Pressure, Cognitive Evaluation and Burnout of Young Doctors

**DOI:** 10.1101/2022.08.08.22278534

**Authors:** Rui-ting Zhang

## Abstract

**Objective:** At present, there are relatively few studies on the burnout of young doctors in China. This study can enrich the relationship between work pressure, cognitive evaluation(benign and stress)and burnout among young doctors, and help to understand the current situation of burnout among young doctors.

**Methods:** A questionnaire survey was conducted among 116 young doctors from different departments to collect data on career motivation, work stress, cognitive evaluation and burnout.

**Result:** (1)The work pressure of young doctors has a predictive effect on burnout.(2)The primary evaluation of young doctors has an independent effect on burnout.(3)The 116 young doctors surveyed in this article are all at a high level of burnout.

**Conclusion:** (1)The work pressure of young doctors has a predictive effect on job burnout. Effective intervention on pay-reward can further reduce the sense of burnout of young doctors and reduce occupational stress.(2)The primary evaluation of young doctors has an independent impact on burnout, and positive cognitive evaluation is beneficial to reduce the level of burnout.

---

In the modern medical environment, medicine has also begun to integrate the elements of spirit and humanity. Treating patients no longer only requires excellent technology, but also requires doctors to care for patients from the humanities. Every doctor should be full of benevolence and responsibility. However, with the continuous increase of clinical needs, the doctor’s task is increasingly arduous, and the psychological pressure on doctors is becoming greater and greater so that they finally fall into a state of “numbness”, which is job burnout. In the process of slowly entering the abyss of job burnout, doctors’ original dreams also slowly turned into a bubble. They will question everything about themselves and separate themselves from the people around them. Anxiety, depression, and despair surround these doctors who rescue the dying and heal the wounded, and they fall into an endless vicious circle. Job burnout not only endangers doctors’ lives but also threatens patients’ life safety. According to the research, the job burnout of clinical doctors is closely related to medical errors, medical accident litigation, and healthcare-related infection rate. Young doctors, as the “special group” of doctors, have to face the complicated doctor-patient relationship, professional title enterprising and self-growth as soon as they leave school. Their job burnout level is relatively high ^[1]^, which may lead to a series of physical and mental disorders, affecting professional performance and continuous commitment. Therefore, the problem of job burnout needs to be paid attention to.

## Literature review

Job burnout, also known as job burnout, was first proposed by American psychologist Herbert Freudenberger in 1974. At present, the most widely-used concept is the three-dimensional definition of job burnout proposed by Maslach and Jackson in 1981, which means “job burnout refers to the emotional exhaustion, depersonalization, and self-achievement produced by people who work in the occupational field where people are the service objects.” Emotional exhaustion is the core of the concept. “ Emotional exhaustion means that a person’s enthusiasm for work has slowly faded and begun to feel tired, unable to work with the same enthusiasm as before. Depersonalization means that a person slowly loses sympathy, becomes indifferent to work, and gradually alienates the work object. A sense of self-achievement means that a person is gradually dissatisfied with his/her performance at work and feels that he/she is incompetent and incompetent^[2]^.

Siegrist, a German physiologist, put forward the work pressure theory of giving back in the late 1990s and elaborated on the formation mechanism and action mechanism of work pressure. Under the condition of an imbalance between contribution and return, employees are prone to a series of diseases, and employees with the tendency of excessive commitment are more seriously affected. Therefore, Siegrist constructed a three-component model with the external assumption of the payoff–return ratio and the internal assumption of the tendency of over-commitment. In this model, both external and internal assumptions have a direct effect on work pressure, and the tendency of over-commitment plays a regulatory role in the effect of the imbalance of pay-out-of-return on work pressure. That is to say, over-commitment can amplify the negative effect of the pay-out-of-return on work pressure^[3]^.

The concept of the cognitive evaluation was first proposed by Lazarus and Folkman. They defined cognitive evaluation as a process of evaluating stress events and all their related aspects, aiming at the significance of stress events for their own health. Lazarus puts forward that primary evaluation and secondary evaluation are two main components of cognitive evaluation, and there is no causal relationship between primary evaluation and secondary evaluation. The primary evaluation includes irrelevant evaluation, benign evaluation, and stress evaluation. Irrelevant evaluation refers to an event having no effect on a person. A benign evaluation is one in which an event has a positive impact on a person. A stressful evaluation is one in which an event negatively affects a person. The stress evaluation is divided into three categories: challenge evaluation, threat evaluation, and important evaluation. Importance evaluations occurred after stress events while challenging and threatening evaluations occurred before stress events. Threatening evaluation refers to the evaluation of the loss that the event may cause, and challenging evaluation refers to the unknown evaluation, such as the growth that the event may bring^[4]^.

In China, the imbalance between doctors’ work effort and reward has been a hot topic of great concern by the government and public opinion in recent years and has also attracted the attention of relevant researchers in the academic research field^[5]^. Only Chu^[6]^and Fang^[7]^have found in the domestic research on the pay-reward imbalance model and job burnout among the medical population that the pay-receive imbalance model can explain the job burnout of nurses to some extent: high pay and low gain have a main effect on emotional exhaustion and depersonalization. More research is needed to determine the relationship between the doctor’s pay-return imbalance and job burnout.

Among the studies on cognitive evaluation and job burnout in China, only Li Xupei found that social support evaluation had no significant prediction on stress response and job burnout except for the positive prediction of support available on job satisfaction^[8]^. The relationship between doctors’ cognitive evaluation and job burnout needs further study ^[9]^.

Therefore, based on the existing research results, the following assumptions are proposed in this paper:

1. Job stress has an impact on job burnout, and a high degree of job stress can predict job burnout.
2. Cognitive evaluation has an impact on job burnout, and stressful cognitive evaluation can predict job burnout.

## Method

### Objects

Using an electronic questionnaire, Beijing Sino-Japanese Friendship Hospital and the First Affiliated Hospital of Inner Mongolia Medical College were selected as the survey sites to investigate the relationship between doctors’ professional motivation, cognitive evaluation, and job burnout among young people (18–44 years old) with low seniority (primary or intermediate title).

### Measuring tool

#### MBI-GS

The Chinese version of the Maslach Job Burnout Scale was adopted with 15 questions in total, which were divided into three dimensions: emotional exhaustion, depersonalization, and self-achievement. Each question had seven scoring options ranging from 0 to 6. The scores of the 15 questions were added up, and the higher the score was, the more serious the job burnout would be.

#### ERI

ERI is a feedback stress work model proposed by Siegrist, a German physiologist, in the late 1990s. It has 20 topics, which are divided into two factors: the pay-to-return ratio (external hypothesis) and the excessive commitment tendency (internal hypothesis). The external hypothesis and the internal hypothesis emphasize that the pay-to-feedback ratio and the excessive commitment tendency have a direct effect on work stress.

#### SAM

SAM was compiled in 1978, with a total of 24 questions, which is divided into two dimensions of primary evaluation and secondary evaluation, of which the primary rating includes three factors of threat evaluation, challenge evaluation, and importance, and the secondary evaluation includes three factors of self-control, others’ control, and uncontrollable. Hudek-Kneevic and Kardum(2000) pointed out that the primary evaluation pointed to immediate reactions with emotional characteristics, while the secondary evaluation mainly considered behavioral intentions and expectations for behavioral efficacy, and more pointed to stress results.

### Statistical method

SPSS20.0 was used for data collection and statistical analysis.

## Result

### General demographic data of study subjects

In the 116 valid questionnaires collected, about two-thirds of the respondents were from tertiary hospitals, and the proportion of young female doctors was relatively large. In the distribution of departments, the Department of Cardiology accounted for the largest proportion. The number of young doctors surveyed who are married and unmarried is mixed.

Their educational background ranges from doctoral degrees to bachelor’s degrees or below, and most of them have a bachelor’s degree or below. The average age of the respondents was 30 years old, and most of them had worked for about seven years.

### Correlation between job burnout and payback ratio, overcommitment tendency, and cognitive evaluation

Return on effort ratio(1.09±0.33); Excess commitment tendency(15.71±2.85); The three dimensions of job burnout—emotional exhaustion (18.44±5.76), depersonalization(13.22±5.22), and self-achievement(23.10±5.85); The three variables in the primary evaluation were threat evaluation(3.06±0.78), challenge evaluation (3.40±0.72), and importance (3.20±0.76). The three variables of secondary evaluation were self-control(3.42±0.74), others’ control(3.32±0.72), and uncontrollable(3.05±0.78).

The results of the correlation analysis (see Table 3) showed that occupation burnout (emotional exhaustion, depersonalization, and self-achievement) showed a medium correlation with the pay-to-return ratio, a medium correlation with the tendency of excessive commitment, a medium correlation with the primary evaluation (threat evaluation, challenge evaluation, and importance), and little correlation with the secondary evaluation (self-control, others’ control, and uncontrollable). Therefore, when discussing the relationship between cognitive evaluation and job burnout, this paper only discusses the primary evaluation.

**Table 1.**
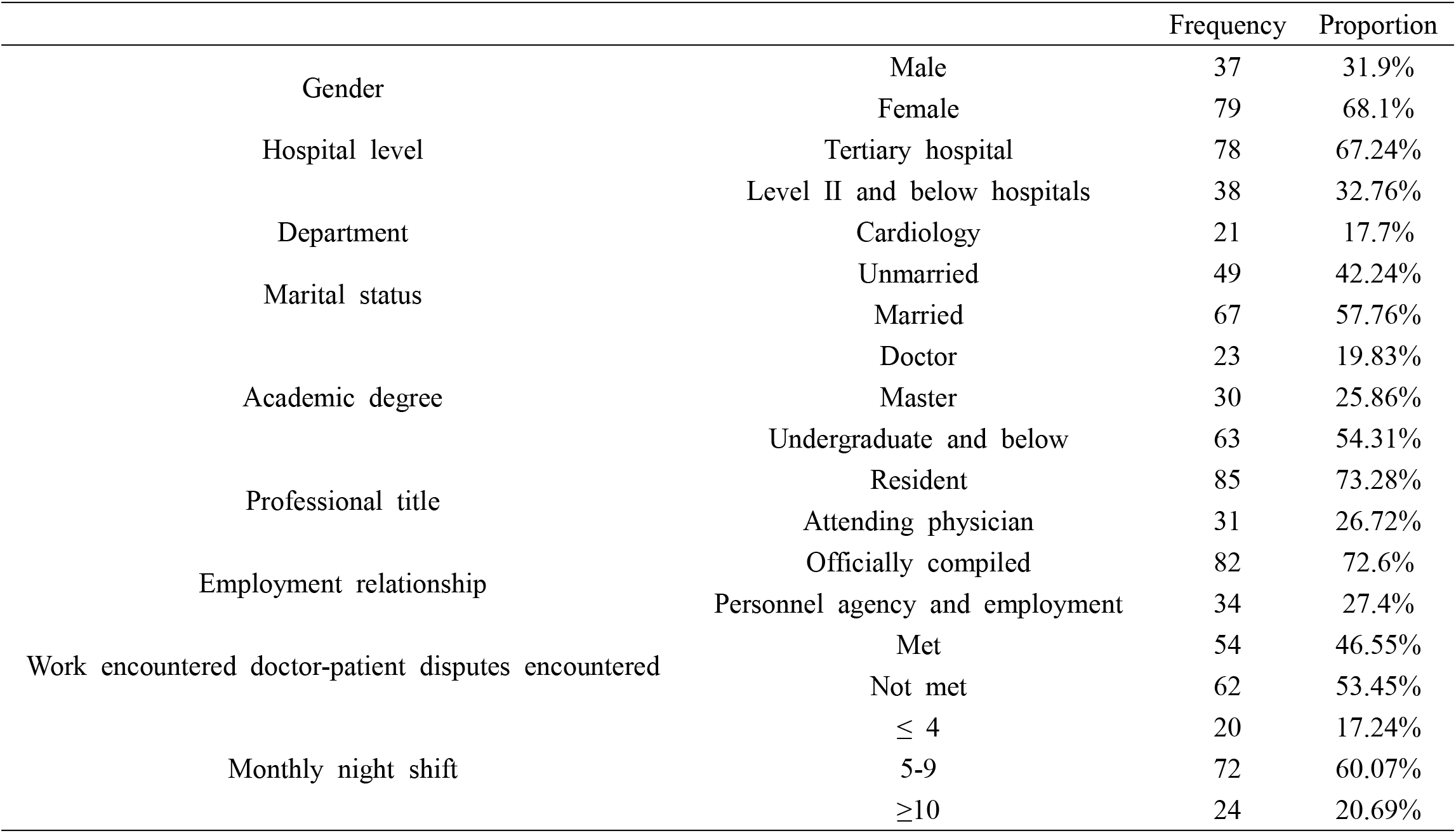
Demographic Information

**Table 2.** Demographic Information

|  | Average | Standard deviation |
| --- | --- | --- |
| Age | 30 | 6.62 |
| Working life | 7.6 | 10.18 |
| Average working hours per day | 9.4 | 0.68 |

**Table 3.** Correlation between variables

|  | 1 | 2 | 3 | 4 | 5 | 6 | 7 | 8 | 9 | 10 | 11 |
| --- | --- | --- | --- | --- | --- | --- | --- | --- | --- | --- | --- |
| 1. Pay-to-return ratio | 1 |  |  |  |  |  |  |  |  |  |  |
| 2. Over-commitment tendency | .54** | 1 |  |  |  |  |  |  |  |  |  |
| 3. Threat assessment | .54** | .51** | 1 |  |  |  |  |  |  |  |  |
| 4. Challenge evaluation | .41** | .13 | .44** | 1 |  |  |  |  |  |  |  |
| 5. Importance | .57** | .48** | .83** | .50** | 1 |  |  |  |  |  |  |
| 6. Self-control | .39** | -.06 | .20* | .76** | .31 | 1 |  |  |  |  |  |
| 7. Control by others | .34** | -.10 | .21* | .70** | .26* | .76** | 1 |  |  |  |  |
| 8. Uncontrollable | .53** | .54** | .85** | .40** | .78* | .19* | .21* | 1 |  |  |  |
| 9. Emotional exhaustion | .55** | .46** | .53** | .06 | .40* | -.08 | .01 | .53* | 1 |  |  |
| 10. De-individualization | .31** | -.30 | .45* | -.02* | .34* | -.14* | -.03* | .46* | .75* | 1 |  |
| 11. Self-achievement | -.04** | -.05* | -.22* | -.49 | -.27 | -.54 | -.49 | -.21 | -.21* | -.13 | 1 |

Note: \* indicates $p < 0.05$ , \*\* indicates $p < 0.01$ , and \*\*\* indicates $p < 0.001$ .

**Table 4-1.** Relationship among work stress, cognitive appraisal and emotional exhaustion

| Step | Independent variable | Normalization $\beta$ | Sig | $\Delta R^2$ |
| --- | --- | --- | --- | --- |
| 1 | Gender | -.128 | .171 | .018 |
|  | Age | .035 | .707 |  |
| 2 | Gender | -.099 | .197 | .344*** |
|  | Age | -.110 | .162 |  |
|  | Pay-for-return ratio | .235 | .011 |  |
|  | Excessive commitment tendency | .445 | .000 |  |
| 3 | Gender | -.041 | .581 | .073** |
|  | Age | -.052 | .500 |  |
|  | Pay-for-return ratio | .198 | .042 |  |
|  | Excessive commitment tendency | .300 | .002 |  |
|  | Threaten | .354 | .000 |  |
|  | Challenge | -.201 | .025 |  |

**Table 4-2.**
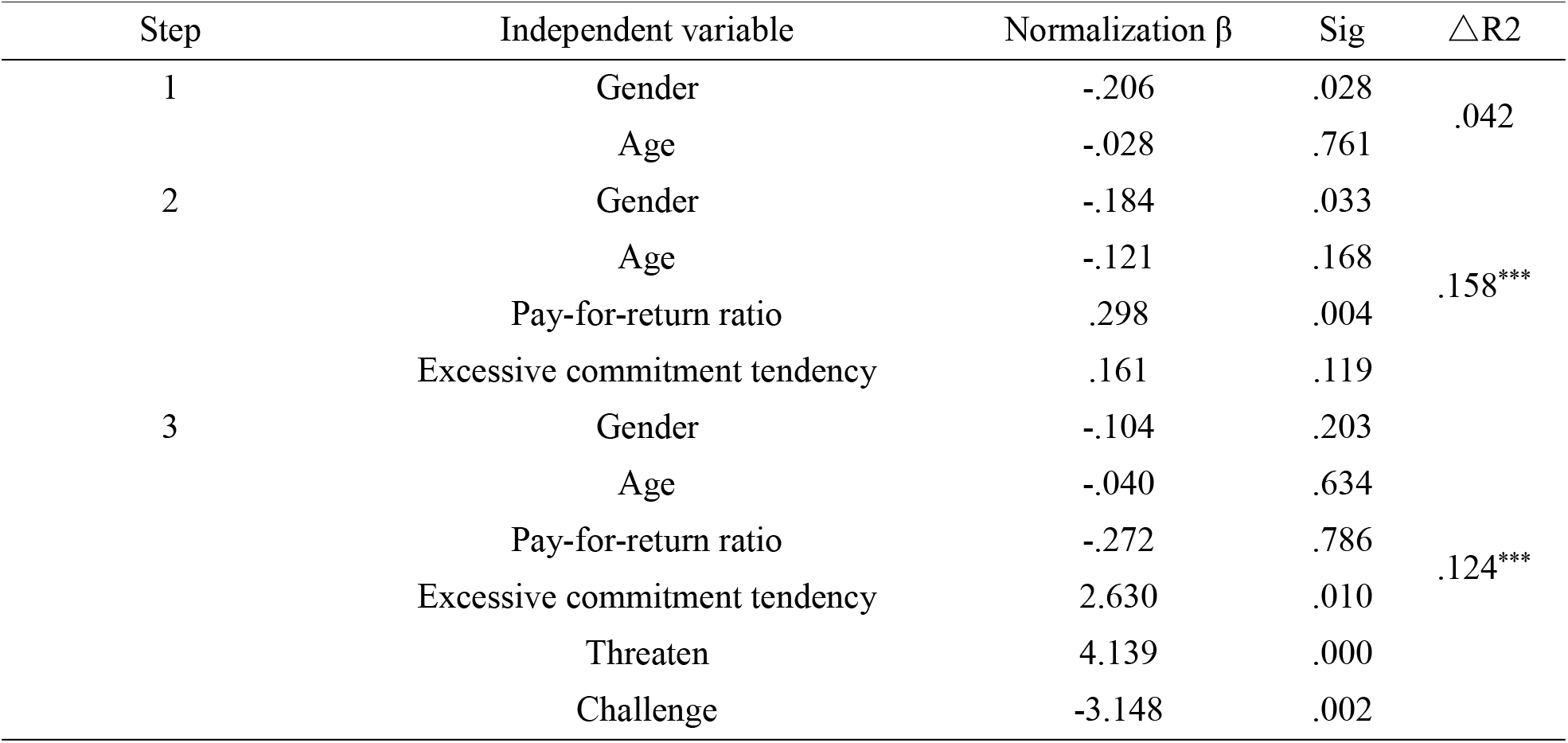
Relationship among work stress, cognitive evaluation and de-individuation

**Table 4-3.**
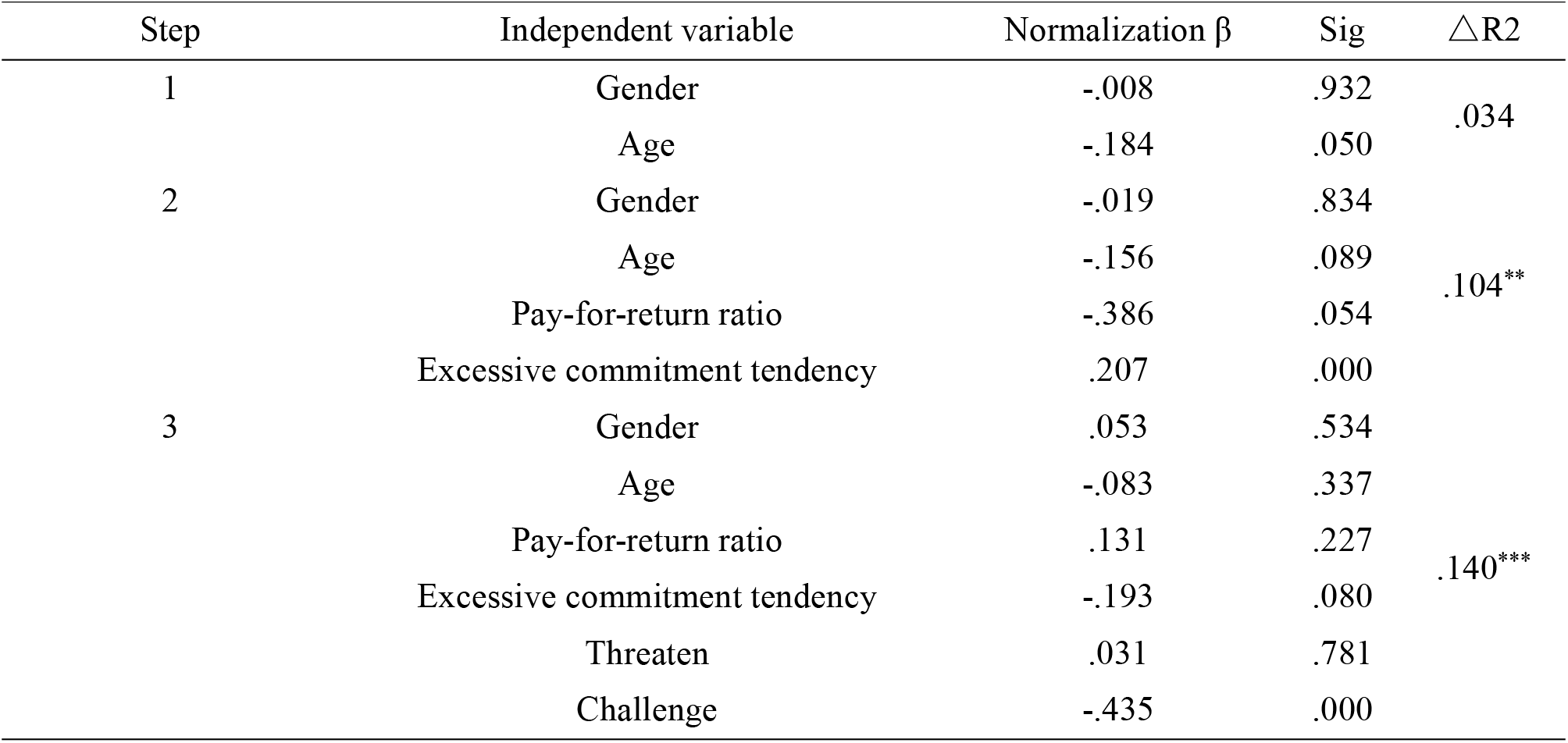
Relationship between work stress, cognitive evaluation and self-achievement

| Step | Independent variable | Normalization $\beta$ | Sig | $\Delta R^2$ |
| --- | --- | --- | --- | --- |
| 1 | Gender | -.008 | .932 | .034 |
|  | Age | -.184 | .050 |  |
| 2 | Gender | -.019 | .834 | .104** |
|  | Age | -.156 | .089 |  |
|  | Pay-for-return ratio | -.386 | .054 |  |
|  | Excessive commitment tendency | .207 | .000 |  |
| 3 | Gender | .053 | .534 | .140*** |
|  | Age | -.083 | .337 |  |
|  | Pay-for-return ratio | .131 | .227 |  |
|  | Excessive commitment tendency | -.193 | .080 |  |
|  | Threaten | .031 | .781 |  |
|  | Challenge | -.435 | .000 |  |

According to the work stress theory of paying back, the factors of paying back and excessive investment have a direct effect on work stress ^[10]^, and the paying-back ratio and excessive commitment tendency have a correlation with job burnout, so it can be concluded that work stress has a predictive effect on job burnout. That is to say, according to the high level of work pressure, individuals can be predicted to be in job burnout, which is in line with hypothesis 1.

### Hierarchical regression analysis

According to the correlation results, the relationship between the three dimensions of job burnout and pay-to-return ratio, excessive commitment tendency, and primary evaluation is further analyzed using hierarchical regression analysis. With emotional exhaustion, depersonalization, and self-achievement as dependent variables, the first level selects gender and age as control variables and eliminates the influence of demographic variables. The second level introduces the pay-to-return ratio and the tendency of excessive commitment to determine their impacts on the three dimensions of job burnout in young doctors. Due to the small sample size, the third level selects the most representative threats and challenges in the primary rating to determine their respective impacts on the three dimensions of young doctors’ job burnout.

After the demographic variables were fixed, the return-to-effort ratio and overcommitment tendency were introduced, and their regression coefficients on emotional exhaustion were 0.235 and 0.445, respectively, and the R2 of the equation increased by 34.4%. After the demographic variables, the pay-to-return ratio and the tendency to overcommit were fixed, threats and challenges were introduced, and the regression coefficients for emotional exhaustion were 0.354 and 0.201, respectively, with the R2 of the equation increased by 7.3%.

After the demographic variables were fixed, the return-to-effort ratio and overcommitment tendency were introduced, and their regression coefficients for depersonalization were 0.298 and 0.161, respectively, and the R2 of the equation increased by 15.8%. After the demographic variables, the pay-to-return ratio and the propensity to overcommit were fixed, threats and challenges were introduced, whose regression coefficients on emotional exhaustion were 4.139 and -3.148, respectively, and the R2 of the equation increased by 12.4%.

After the demographic variables were fixed, the return-to-effort ratio and overcommitment tendency were introduced, and their regression coefficients on emotional exhaustion were -0.386 and 0.207, respectively, and the R2 of the equation increased by 10.4%. After the demographic variables, the pay-to-return ratio and the tendency to overcommit were fixed, threats and challenges were introduced, whose regression coefficients on emotional exhaustion were 0.031 and 0.435, respectively, and the R2 of the equation increased by 14%.

The comprehensive three-table analysis showed that the pay-return ratio, over-commitment tendency, and primary evaluation had independent effects on the three dimensions of young doctors’ occupational burnout. In addition, the pay-return imbalance and over-commitment tendency had significant effects on young doctors’ occupational burnout and had a strong explanation for it.

According to the work stress theory of paying feedback, the factors of paying feedback and excessive input have a direct effect on work stress^[11]^. Therefore, according to the hierarchical regression result, assuming that once verified again, work stress has an effect on job burnout and a high degree of job stress can predict job burnout.

The stressful evaluation in the primary rating is divided into three parts: challenge evaluation, threat evaluation, and importance evaluation. Therefore, according to the hierarchical regression result, hypothesis two is also verified. The cognitive evaluation has an effect on job burnout, and the stressful cognitive evaluation predicts job burnout.

## Discuss

### Analysis of the current situation of pay-to-return ratio, over-commitment tendency, cognitive evaluation, and job burnout of young doctors

According to the questionnaire statistics, young doctors in the scope of the survey have a high degree of job burnout. The higher the scale score is, the more serious the job burnout becomes. The three dimensions of sample emotional exhaustion(18.44±5.76), depersonalization(13.22±5.22) and self-achievement (23.10±5.85) obtained in this study have high scores, which indicates that young doctors in the scope of the survey have a high degree of job burnout.

According to the recycling questionnaire, the pay-to-return ratio of all samples was positive, the mean and standard deviation of the pay-to-return ratio were1.09±0.33, and the mean and standard deviation of the over-commitment tendency were15.71±2.85. It can be seen that the pay-to-return imbalance faced by young doctors in the survey widely existed, and the pay-to-return ratio was generally greater than the return. Workload overload and effort-reward imbalance were significant risk factors for depression among medical staff^[10]^.From the results of the correlation test and hierarchical regression analysis, there was a moderate correlation between the pay-to-return ratio and the tendency of excessive commitment and the three dimensions of job burnout, and they positively affected the emotional exhaustion and depersonalization and negatively affected the self-achievement sense. That is to say, the more young doctors pay, the more tired they feel about their work. They gradually lose their original enthusiasm for work and begin to feel dissatisfied with their current situation. Eventually, they fall into a vicious circle of self-pity and a continuous decline in work efficiency. According to the work stress theory of paying back, the factors of paying back and excessive investment have a direct effect on work stress^[11]^, and the paying-back ratio and excessive commitment tendency have a correlation with job burnout, so it can be concluded that work stress has a predictive effect on job burnout. That is, according to the high degree of work pressure, individuals can be predicted to be experiencing job burnout, which also provides ideas for the intervention of job burnout.

From the results of hierarchical regression, it can be seen that cognitive evaluation, especially primary evaluation, has an independent impact on the three dimensions of job burnout and can also affect the result of stress, but the specific relationship needs further study.

### Compared with relevant domestic surveys

According to a study conducted by the Hong Kong Polytechnic University over the past 18 years, the job burnout rate of Chinese doctors is actually as high as 66.5%–87.8%. In other words, more than two-thirds of Chinese doctors are experiencing job burnout, and 12.1%–25.4% of them suffer from serious job burnout ^[12]^. The 116 young doctors surveyed in this paper are all at a high level of job burnout. Although they cannot represent the national level, we can deduce the probability of job burnout among young doctors at present. Long-term high-level job burnout will lead to physical and mental fatigue of young doctors. Their concern for everything will decline, and they will even ignore their future and that of the hospital ^[13]^, and they will gradually become indifferent to all the people and things around them, which may eventually lead to irreparable consequences.

The pay-to-return ratios in this paper were all positive, which was consistent with the survey results by Fang Yanyu et al ^[11]^, and indicated that the pay-to-return ratio of young doctors was greater than the return ratio. This may be related to a lack of experience and inexperience, or it may be related to the situation of patients in the hospital.

In summary, the comparison between this survey and related surveys in China shows that young doctors are at a high risk of high occupational burnout, which is consistent with the latest survey results in China. The young and middle-aged doctors in this survey generally paid more than they received in return. From the results of correlation analysis, it could be seen that the imbalance of payment reporting positively predicted job burnout, which was consistent with the known survey of nurses^[6,7,11]^. Therefore, occupational stress on job burnout also exists in the prediction, which also verifies the Fang Yanyu et al^[11]^. research results. According to the stratified regression results of this survey, it can be concluded that cognitive evaluation has an independent influence on job burnout from occupational stress, but it does not reach the intermediary effect in the existing research results.

### Suggestions on coping with high job burnout

For a long time in high pay and high load under work pressure, health is bound to have a long-term negative impact, so medical institutions should design a scientific and reasonable salary system and performance evaluation index^[14]^to reflect the technical labor value of medical personnel, so that young doctors feel their pay in return, thus improving job satisfaction. At the same time, some policies should be promulgated to protect the reasonable working hours of doctors^[15]^and reduce the dedication of young doctors to achieve the purpose of reducing job burnout.

According to the stratification results, it can be seen that the excessive pay tendency of young doctors also affects occupational burnout. Therefore, in the education of medical colleges and universities, we should strengthen the personality education of medical students to correct their excessive pay behavior so as to reduce their vulnerability to occupational depletion.

In the face of pressure, a person’s evaluation of pressure largely determines the response and performance in dealing with pressure, and positive cognitive evaluation will not only directly affect the level of pressure but also have an independent impact on job burnout, so young doctors can practice more positive pressure evaluation in daily life, and gradually form a positive response habit, master the skills of doctor-patient communication, avoid doctor-patient disputes and other conflicts, and intervene when job burnout is budding, to improve work happiness. At the same time, medical colleges and hospitals can also take some measures to help medical students and young doctors to establish a positive cognitive evaluation, such as organizing some youth league activities and class activities for group psychological intervention, to help medical students and young doctors to better cope with professional pressure.

This study has certain limitations. In this study, convenient sampling was adopted, but the sample size was small, so the results were difficult to extrapolate. Convenience sampling was adopted in the questionnaire survey, which was less representative. A cross-sectional survey was adopted, and no longitudinal survey was conducted. The specific connection of each variable needs to be further explored in the future. In the future, prospective longitudinal research can be conducted to fully analyze the relationship between cognitive evaluation and job burnout as well as other influencing factors of job burnout.

## Data Availability

All data produced in the present study are available upon reasonable request to the authors.
All data produced in the present work are contained in the manuscript.

